# Vaccine breakthrough and the invasion dynamics of SARS-CoV-2 variants

**DOI:** 10.1101/2021.12.13.21267725

**Authors:** Chadi M. Saad-Roy, Simon A. Levin, Julia R. Gog, Jeremy Farrar, Caroline E. Wagner, C. Jessica E. Metcalf, Bryan T. Grenfell

## Abstract

Vaccination provides a powerful tool for mitigating and controlling the COVID-19 pandemic. However, a number of factors reduce these potential benefits. The first problem arises from heterogeneities in vaccine supply and uptake: from global inequities in vaccine distribution, to local variations in uptake derived from vaccine hesitancy. The second complexity is biological: though several COVID-19 vaccines offer substantial protection against infection and disease, ‘breakthrough’ reinfection of vaccinees (and subsequent retransmission from these individuals) can occur, driven especially by new viral variants. Here, using a simple epidemiological model, we show that the combination of infection of remaining susceptible individuals and breakthrough infections of vaccinees can have significant effects in promoting infection of invading variants, even when vaccination rates are high and onward transmission from vaccinees relatively weak. Elaborations of the model show how heterogeneities in immunity and mixing between vaccinated and unvaccinated sub-populations modulate these effects, underlining the importance of quantifying these variables. Overall, our results indicate that high vaccination coverage still leaves no room for complacency if variants are circulating that can elude immunity, even if this happens at very low rates.

## Introduction

The remarkably rapid development of effective vaccines against SARS-CoV-2 opens a way for sustainable control of the pandemic. At best, vaccines achieve their power by combining direct protection of vaccinees against infection and serious disease with indirect ‘herd’ protection of susceptible individuals by reduction of transmission. Though this combination can eliminate infection on local or broader scales^1^, two sets of forces can compromise this ideal.

First, vaccination rates may not be sufficiently high to interrupt transmission. In the context of the COVID-19 pandemic, inequities in global vaccine distribution^2^, as well as vaccine hesitancy^3^ are significant brakes on vaccination rates at different scales. Second, the impact of the vaccine may be reduced by biological factors, notably immunological complexities^4–7^ and viral evolution^6–9^. We can illustrate these forces by reference to the simplest, most optimistic case where vaccination and natural infection provide lifelong immunity against reinfection. This leads to the standard SIR (Susceptible-Infected-Recovered) epidemic model, wherein transmission can be interrupted in a well-mixed population by immunizing a proportion

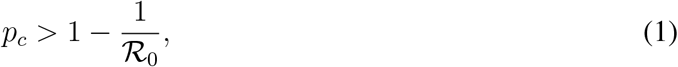

where ℛ_0_ is the basic reproduction ratio of infection (the number of secondary cases caused by one infectious individual in an otherwise fully-susceptible population^10^); note that the proportion immunized (*p*_*c*_) will generally be lower than the proportion *vaccinated*, to allow for vaccine efficacy. In practice, the strong and prolonged immunity implicit in this model is only seen in a minority of pathogens^11^. Gaps in the ability of natural or vaccinal immunity to block infection and subsequent transmission can derive from a combination of immune evasion by pathogens and the evolution of immune escape (classically seen in ‘antigenic drift’ by seasonal influenza^12–14^, and recently identified in the seasonal coronavirus 229E^15^). SARS-CoV-2 embodies both these pathogen strategies, as well as the evolution of increased transmissibility exhibited for example by the delta variant^16^.

The dynamics of imperfect immunity are captured by epidemiological models from the SIRS family^10, 11^. In particular, the SIR(S) model gives a parsimonious and flexible accounting for the potential impact of imperfect and transient immunity^5, 11^. Two SIR(S) parameters capture current uncertainties about the prevalence and effects of COVID-19 vaccine breakthrough infections. First, *ϵ* is the relative susceptibility to breakthrough infection: *ϵ* = 0 equates to perfect and lifelong transmission-blocking immunity (SIR dynamics), whilst *ϵ* = 1 generates SIRS dynamics: a return to complete susceptibility after immunity has waned^5^ (note that we do not explicitly analyze the important processes of waning and boosting of immunity in this paper). The second key parameter, *α*, is the relative transmission rate of secondarily-infected individuals, compared to that following infection of naive susceptibles. The parameters *ϵ* and *α* therefore combine to modulate the net transmission rate from breakthrough infections.

### Estimating ϵ and α

In the early stages of the pandemic (especially pre-vaccination), a lack of testing and longitudinal infection data made estimation of these important parameters nigh on impossible, especially given changes in vaccine effectiveness against infection. Recently, however, a number of studies have started to address these questions^17–22^. In particular, Pouwels *et al*.^20^ *follows large groups of randomly tested individuals to calculate a recent Vaccine Effectiveness (V E*) against infection of around 70% in the UK; this figure is bracketed by other recent estimates^17–19^, ^21^. In the Supplementary Information (section 4), we show that relative susceptibility in the simple model of the next section equates approximately to *ϵ* = 1 − *V E*, when incidence is low at the start (and end) of outbreaks. Furthermore, we demonstrate that higher incidence of infection magnifies estimates of *V E*; thus, if anything, 1 − *V E* is a lower bound for the breakthrough susceptibility, *ϵ*. Given the range of uncertainties, plus the possibility of future escape variants, we explore a range of values for *ϵ* below. Clearly, however, recent estimates of *V E* indicate that *ϵ* is not close to zero, consistent with a degree of susceptibility to vaccine breakthrough.

Recent estimates of the relative transmissibility, *α*, of the delta strain in vaccinated individuals indicates high transmission, comparable to viral shedding in naive, susceptible individuals^23^; however, the duration of breakthrough shedding may be shorter^24^. In line with previous work^5–7^, we initially make the pessimistic assumption that *α* = 1, then explore the impact of lower values.

In the next section, we begin with a simple model for the invasion of a new strain into a partially-vaccinated population, exploring how the interaction of primary and secondary infection shapes the resulting outbreak. The initial motivational model assumes a well-mixed, immunologically-homogeneous population. In subsequent sections, we consider how a number of biological refinements tune our results. In particular, we explore the impact of immunological heterogeneity, via polarized vaccinal immunity^25^, where some individuals are completely susceptible to breakthrough infections. We also explore the impact of population-level transmission heterogeneities on our results, notably the social separation between vaccinated and susceptible individuals seen in some settings ^26^.

### A homogeneous variant invasion model

#### Basic framework

We explore the interaction between primary population susceptibility and vaccine breakthrough using the following simple, relatively optimistic scenario. Assume that a country well supplied with well-matched vaccines has achieved vaccinal herd immunity against the prevailing strain. Again optimistically, assume SIR dynamics and a reproduction ratio ℛ_0,*ϵ*_ for this endemic variant. Herd immunity to eliminate the current variant is then achieved by reducing the fully susceptible proportion of the population to or below *S* = 1*/*ℛ_0,*ϵ*_.

A new variant now invades the population at a low initial prevalence *I*(0). The variant has a reproduction ratio ℛ_0,*I*_ for fully susceptible individuals. Initially, we assume ℛ_0,*I*_ *>* ℛ_0,*ϵ*_; this assumption is relaxed below. Finally, we assume that the new variant causes some breakthrough infection from a ‘leaky’ vaccine, such that the reproduction ratio of the new variant on vaccinated individuals (*i*.*e*., the effective reproduction number in a fully vaccinated population) is 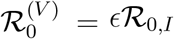, where 0 ≤ *ϵ* ≤ 1 is the proportional reduction in susceptibility for vaccinees (and the resulting relative onward transmission, *α* = 1). Finally, *γ* is the recovery rate of infected individuals who move into the recovered class, *R*. The dynamics of the ensuing outbreak are then controlled by the following equations:

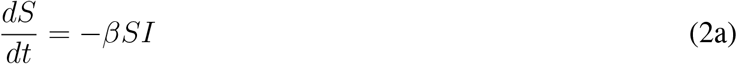

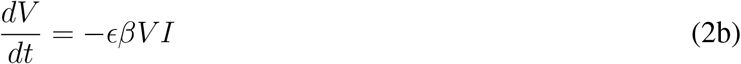

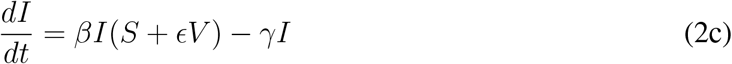

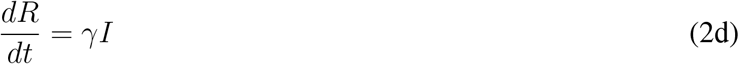

We neglect host vital dynamics, model the outbreak of the new variant as a simple epidemic, and initially consider a homogeneously-mixed population. For simplicity, the previous variant is assumed to be locally eliminated. Furthermore, we make the optimistic assumption that natural infection is fully immunizing. *S* are susceptible individuals (all state variables are proportions of the total population) and *V* those vaccinated against the previous variant at the start of the outbreak. To focus on the ensuing epidemic dynamics, we assume that further vaccinations do not occur during the outbreak.

The baseline infection rate of susceptibles is *β* and the recovery rate from infection is *γ*; The associated basic and effective reproduction ratios on susceptibles are *R*_0,*I*_ = *β/γ* and 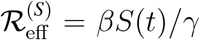 respectively. The corresponding infection rate of vaccinees is *ϵβ* and we assume the same recovery rate *γ*. Thus the vaccinee reproduction ratios are 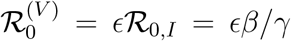 and 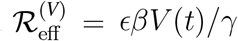.

#### Model dynamics

The new variant invades at time zero, at a low prevalence, *I*(0) = 10^−6^ (qualitative results are not sensitive to this exact value); the initial susceptible proportion is *S*(0) = 1*/*ℛ_0,*ϵ*_. Initially, we assume that vaccination does not completely immunize anyone against the new variant, so that *R*(0) = 0 (this pessimistic assumption is relaxed below). Consequently, the vaccinated proportion: *V* (0) = 1 − *S*(0) − *I*(0) − *R*(0) dominates the initial population. Finally, we set the average duration of infection to 1*/γ* = 1 week.

We explore the resulting invasion dynamics of the new variant by calculating the final size of the simple epidemic (see Eq. (S1), Supplementary Information). Figure 1A shows the final size as a function of the vaccinee reproduction ratio 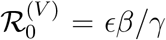. As expected, for zero vaccine breakthrough (*ϵ* = 0), the epidemic is small and limited to primary susceptibles. As 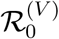 increases, the final size rises monotonically as more vaccinees become infected. Though this qualitative pattern is intuitive, the rapid increase in the epidemic seen in Figure 1A (and the associated inset epidemic plot) is surprising – the epidemic reaches almost a third of the population even though the associated vaccinee reproduction ratio is only around 0.5.

**Figure 1:**
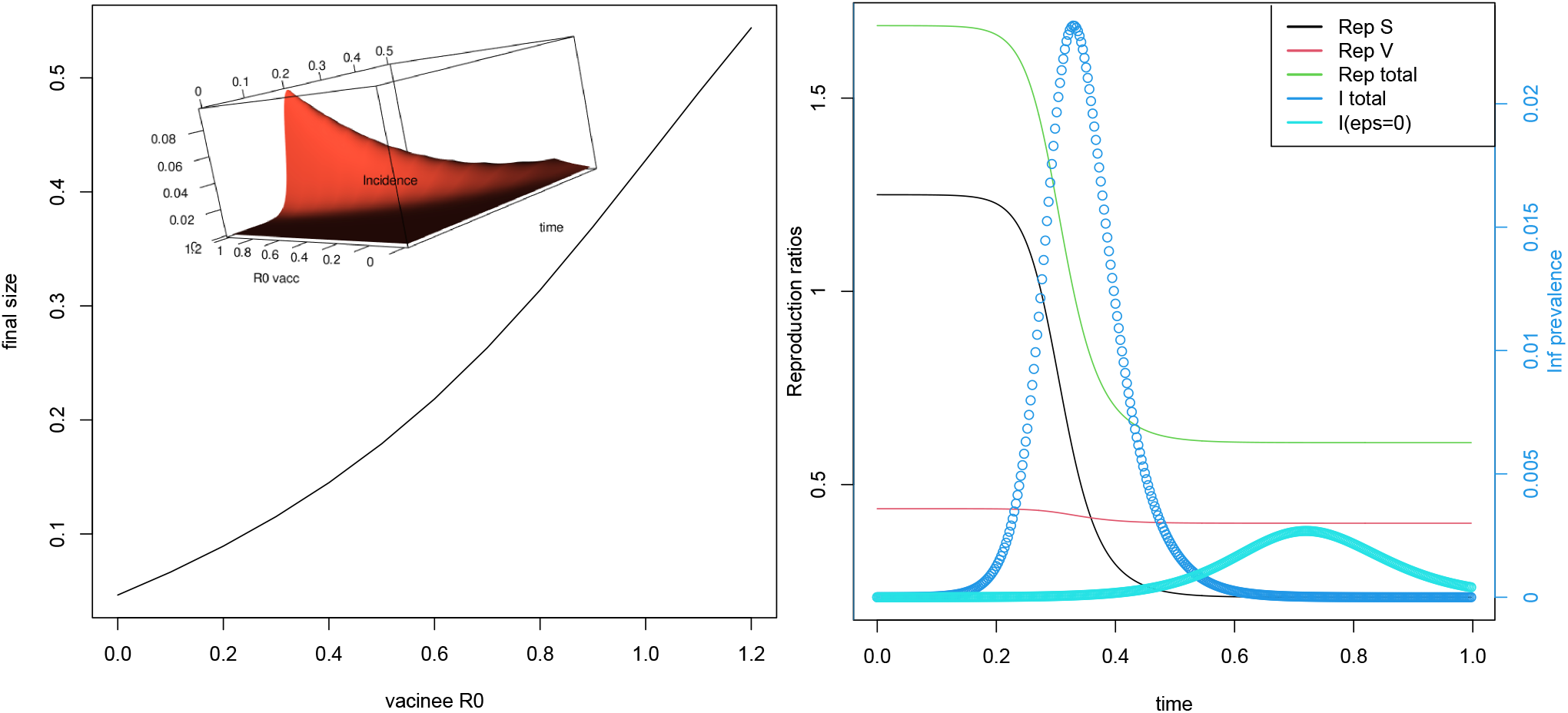
Invasion dynamics of the basic model: Equations 2. *A* (Left), final size with inset epidemic as a function of 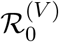, *B* (Right) Epidemic and associated reproduction ratios for *ϵ* = 0.05. The pale blue curve is the equivalent epidemic for *ϵ* = 0. Note that “Rep S” denotes 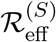, “Rep V” denotes 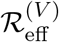, and “Rep total” denotes 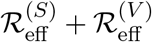.

The explanation for this rapid rise in the final size becomes clear if we consider the effective reproduction ratio of infection for the invading variant. For infection to increase in Equations (1), we require

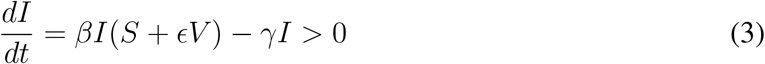

Hence

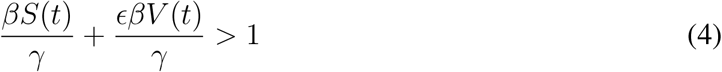

As defined above, the two terms on the left of the inequality are the effective reproduction ratios for susceptible and vaccinated individuals respectively, i.e.,

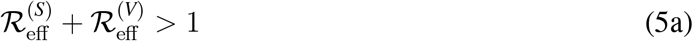

The total reproduction ratio of infection is thus the sum of transmission in the two population components. The resulting outbreak dynamics are illustrated in the simulation in Figure 1B, where the invading reproduction ratio, ℛ_0,*I*_ = 10 and *ϵ* = 0.05. This leads to an initial epidemic in the susceptible population which drives infection into the vaccinated group, even though their reproduction ratio 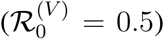 is significantly less than unity. Because there are so many vaccinated individuals, this leads to a larger and more violent epidemic (dark blue curve in Figure 1B) than would be observed if there were no vaccine breakthrough infections (*ϵ* = 0, light blue curve).

We can further dissect the large impact of a relatively low breakthrough infection rate on the susceptible and vaccinated populations by separating the infected class into infections from primary, naive susceptibles (*I*_*S*_) and from vaccinees (*I*_*V*_). Correspondingly, *R*_*S*_ and *R*_*V*_ are recovered individuals from the susceptible and vaccinated classes. The resulting epidemic model is:

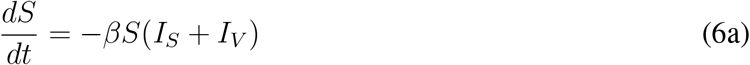

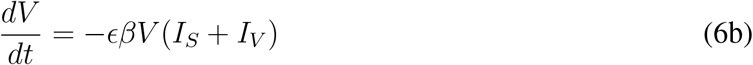

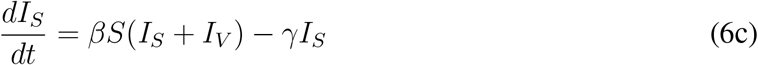

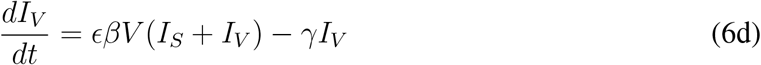

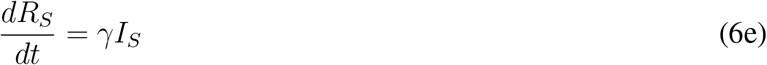

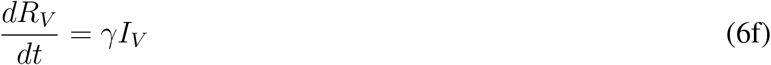

Figure 2 uses this model to repeat the final size calculation, split into susceptible and vaccinated epidemics and across a range of breakthrough proportions, *ϵ*, of the invading variant. Focusing first on proportion infected of total population (Figure 2A), these results bear out Figure 1A, in that both susceptible and vaccinated final epidemic sizes can be magnified by their interactions even if the reproduction ratio of infection of vaccinees is less than unity (area to the left of the red vertical line). This pattern even holds when the invading reproduction ratio, *R*_0,*I*_ is less than that of the previous variant (ℛ_0,*I*_ = 6, 7) as long as *ϵ* is sufficiently high.

**Figure 2:**
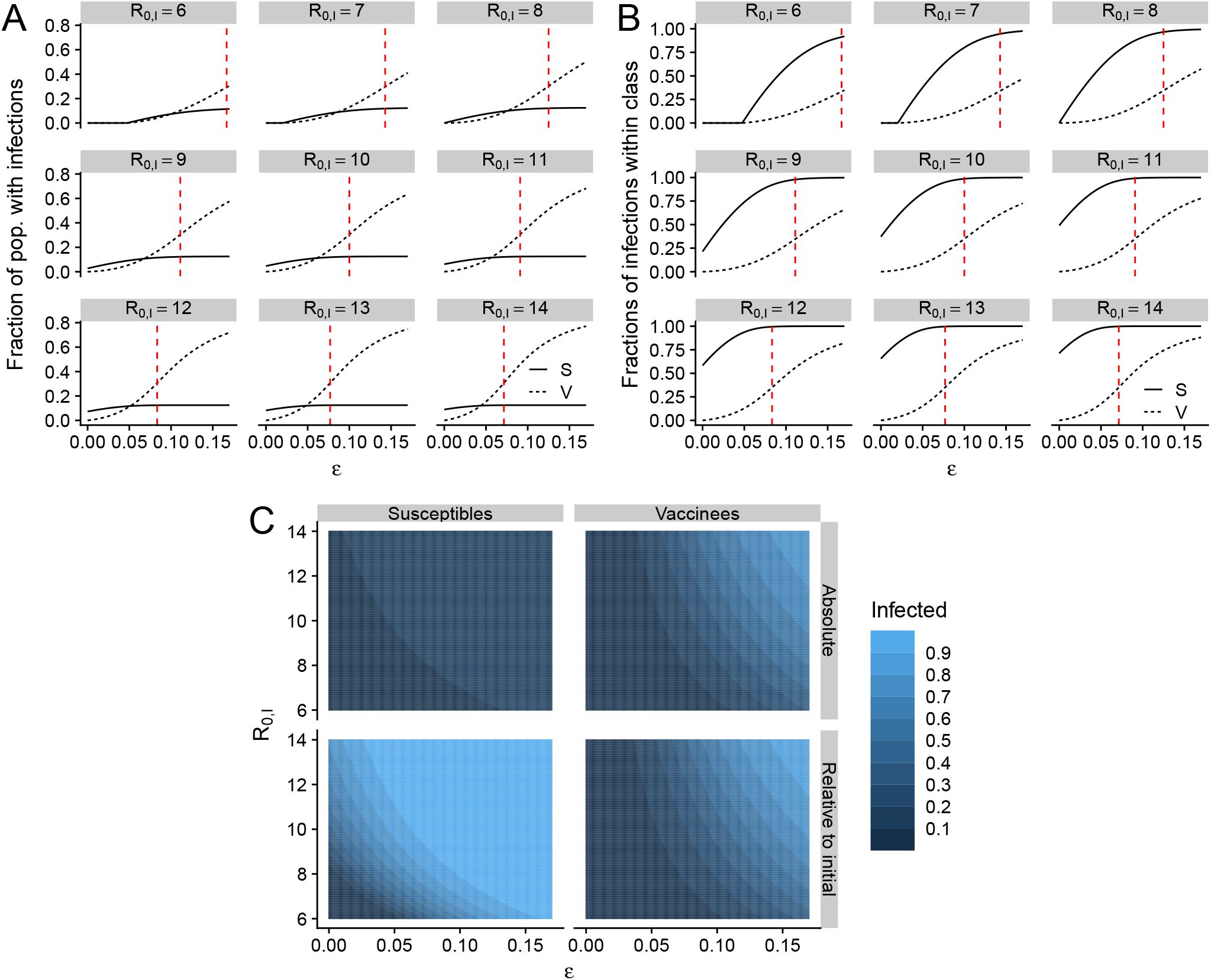
Illustrations of final size relations in the simple epidemic model. (*A*) The final fractions of the total population that become infected among susceptibles (solid line) and vaccinees (dashed lines). (*B*) The relative fraction of initially susceptible or vaccinated individuals that become infected during the epidemic. In both (*A*) and (*B*), each sub-panel depicts a different invasion ℛ_0,*I*_ and the vertical dashed red line is the value of *ϵ* for which *ϵ*ℛ_0,*I*_ = 1. (*C*) The absolute (*top row*) and relative to initial fractions (*bottom row*) of susceptibles (*left column*) and vaccinees (*right column*) that are infected during the epidemic as functions of *ϵ* and ℛ_0,*I*_.

Given the ability of current COVID-19 vaccines to reduce clinical severity, the division between infection of primary susceptibles and vaccinees also stresses that much of the increase in total cases in Figure 2A will hopefully be mild infections. However, another perspective on this result is illustrated in Figure 2B, where we plot the associated *proportion* of initial susceptibles and vaccinees infected. Here, the proportion of susceptibles infected via the vaccinal epidemic is much larger as *ϵ* increases, illustrating the impact of the breakthrough epidemic.

In Figure 2C, we present these fractions and proportions of infected individuals across a range of values for the reproduction ratio of the invading strain. In a largely vaccinated population, a variant with a slight increase in *ϵ* leads to larger epidemics among both the susceptibles and the vaccinees than those caused by more transmissible variants (compare horizontal and vertical axes). In addition, the worst epidemics occur when a variant has a combination of higher transmissibility and vaccine escape (top right of each panel). As before, these results are due to the feedback between infected vaccinees and susceptibles enabled by transmissible breakthrough infections.

### Immunological uncertainties

#### Transmissibility of breakthrough infections

So far, in line with recent evidence that infectious break-through infections may be relatively transmissible^23, 27^, we have made the conservative assumption that such infections are as transmissible as those in fully susceptible individuals. In the Supplementary Information, we relax this assumption and instead assume that the relative transmission of breakthrough infections is 0 ≤ *α* ≤ 1. We obtain a corresponding final size relation for this expanded model (see Eq. (S7)), and in Figure S1 present final size values across ranges of *α* and *ϵ*. Intuitively, our results are qualitatively similar across values of *α*, though with dampened epidemic sizes in both susceptibles and vaccinees if breakthrough infections are substantially less transmissible.

#### Dynamics of susceptibility in vaccinees

Our basic model makes the simplest (relatively pessimistic) assumption that all vaccinated individuals are equally susceptible to breakthrough infection; this corresponds to a leaky vaccine, controlled by the susceptibility parameter, *ϵ*. A more optimistic scenario would be a mixed history-polarized immunity model^28^, where a proportion *ϵ*_2_ of vaccinees are potentially fully immune to breakthrough; thus, a proportion 1 − *ϵ*_2_ of vaccinees have a reduced susceptibility to breakthrough during the invading outbreak. As before, breakthrough infection in the reduced-susceptibility group (1 − *ϵ*_2_)*V* is controlled by the proportional parameter *ϵ*_1_. Final size calculations for this refined model are described in Eqs. (S8), Supplementary Information.

In practice, we still have little idea for SARS-CoV-2 of the immune heterogeneity determined by the balance between *ϵ*_1_ and *ϵ*_2_; Figure 3 thus explores a range of values of these parameters. As complete immunity after vaccination (*ϵ*_2_) increases (denoting increasingly optimistic scenarios for vaccinal immunity), the fraction of susceptible individuals that are infected in the epidemic decreases, especially for larger values of *ϵ*_1_ (vertical transects, bottom-left panel). For large enough *ϵ*_1_, infections among vaccinees markedly decrease as *ϵ*_2_ increases (vertical transects, bottom-right panel). Therefore, it is critical to determine the exact immunological status of hosts that encounter breakthrough infections, in order to properly estimate the relative contributions of *ϵ*_1_ and *ϵ*_2_ to invasion dynamics.

**Figure 3:**
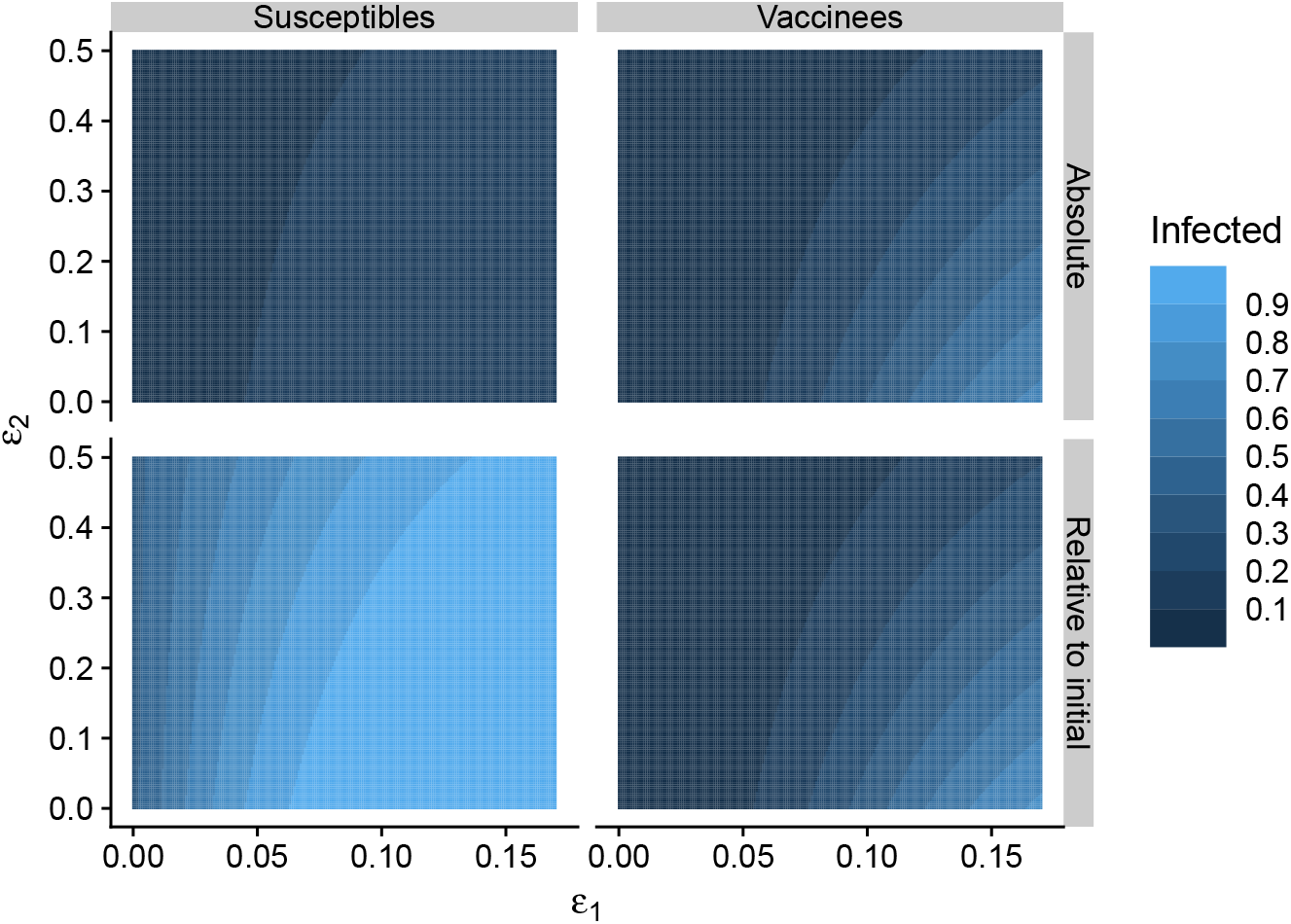
The impact of the nature of immunity on epidemic sizes. The absolute and relative final sizes are determined as a function of the two immunity parameters *ϵ*_1_ and *ϵ*_2_, where *ϵ*_2_ denotes the fraction of vaccinated individuals with total immunity (*i.e*. cannot get infected), and *ϵ*_1_ is the reduction in susceptibility imparted by the presence of vaccinal immunity in the 1 − *ϵ*_2_ fraction of ‘susceptible’ vaccinated individuals. The sub-panels for this figure are as in Figure 2C.

### Impact of transmission heterogeneities

Transmission heterogeneities can have significant impacts on epidemic dynamics and prospects of control^5, 10^. We focus first on an important heterogeneity in the current pandemic, which may be particularly salient for the dynamics of vaccine breakthrough. Specifically, we consider a simple model for imperfect mixing between vaccinated and susceptible populations, due to underlying spatial or social heterogeneities. We achieve this by reducing transmission between susceptible and vaccinated groups by a factor *κ*, 0 *< κ* ≤ 1 (*κ* = 1 recovers the homogeneous model). This leads to the following modification of Equations (5).

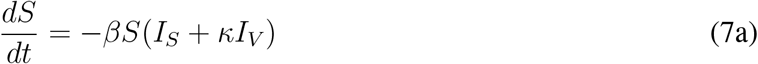

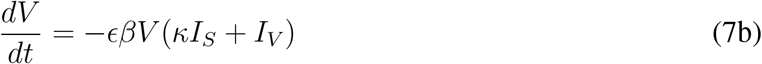

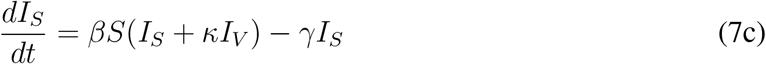

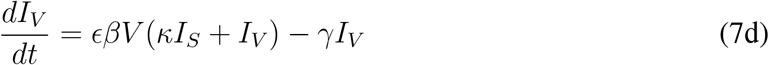

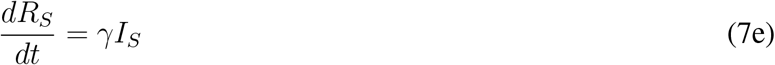

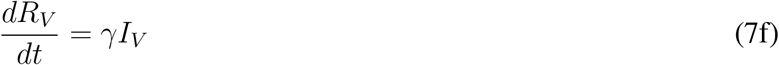

The final size calculation for this model is set out in Eqs. (S10) (Supplementary Information) and plotted as a function of *ϵ* and *κ*. Intuitively, Figure 4 shows that reducing across-group transmission can mitigate the impact of the dual epidemic, reducing number of infections in both susceptible and vaccinated groups. This is partly because of the heterogeneity but also because reducing *κ* lowers average transmission. In the Supplementary Information (see Eqs. (S11) and Fig. S2), we analyze a model which scales *κ* such that average *R*_0_ is constant; this generates much more subtle effects on the dual epidemic. For example, if vaccinees alone cannot sustain an epidemic, then heterogeneity in transmission tends to increase the number of infections among susceptible individuals. On the other hand, if transmission is sustained among vaccinees, then heterogeneity may lead to fewer infections among the susceptibles (Fig. S2).

**Figure 4:**
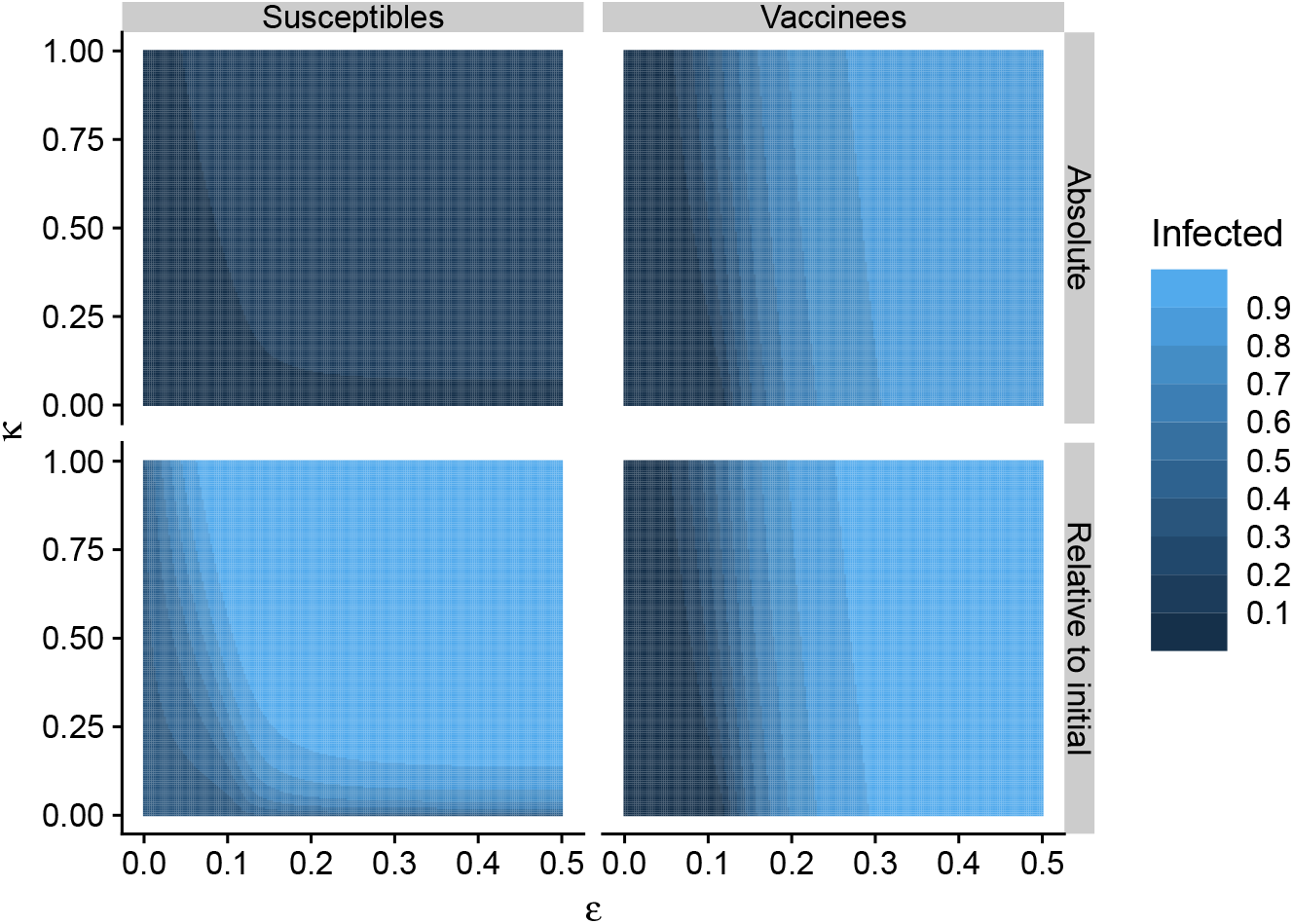
The impact of simple heterogeneity in transmission on epidemic sizes. We assume 0 *< κ* ≤ 1 denotes strength of interactions between the susceptibles and vaccinees, and examine the interaction of *κ* with *ϵ*. In this figure, all sub-panels are as in Figure 2C.

## Discussion

The consequences of the dynamics of vaccine breakthrough by invading viral variants are emerging as a vital issue for the future of the COVID-19 pandemic. Here, we use simple models to explore the potential interaction between outbreaks in susceptible and vaccinated sub-populations. We show that a susceptible epidemic can potentiate vaccine breakthrough even when the reproduction ratio for vaccinees is less than unity. In turn, transmission from a large vaccinated population can drive the susceptible epidemic. We show that this potentiation arises because the reproduction ratio of the invading variant is the sum of transmission in the susceptible and vaccinated classes. By way of an ecological analogy, the invading variant is acting as a ‘generalist predator’^29^ with growth rate driven by two sources of ‘prey’. We stress that these interactions are likely to appear in more complex models for imperfect vaccinal immunity (eg,^5^); our aim here was to explore the interaction explicitly with simple models.

To characterize these dynamics further, we need to measure the susceptibility to vaccine breakthrough. As reviewed in the Introduction, there are still considerable uncertainties in the estimation of *ϵ*; however, it does appear as though *ϵ* is markedly larger than zero for the delta variant^20^. The results in Figure 2 give a qualitative feel for the impact of this parameter: if *ϵ* is small, for example if *ϵ <* 0.01, there will be a small impact of the interacting epidemics. However, if there is more breakthrough (higher *ϵ*) for current or future variants, the effect could be much stronger, especially if the invader has an increased reproduction ratio over current infections. On the latter point, Figure 2 underlines that reduction of an invading ℛ_0_ by non-pharmaceutical interventions or frequent testing is a crucial adjunct to mass vaccination.

The onward infection parameter after breakthrough, *α*, is also an important parameter. In line with recent estimates^23^, we take the conservative approach of setting *α* to unity. We then show that reducing *α* mitigates the interaction of breakthrough with the overall epidemic; note, however, that *α* could in principle *increase* above unity for future variants, worsening the impact of vaccine breakthrough.

Our modeling makes a number of simplifying assumptions, some of which prompt areas for future work. These can be usefully be categorized as pessimistic versus optimistic in terms of projected infection rates. Immunologically, our assumption of strongly immunizing (SIR) dynamics for natural infection is optimistic; weaker immunity would increase the interaction between transmission from vaccinal versus natural breakthrough. In terms of immunological heterogeneity, we began by assuming uniform ‘history-based’ immunity across all vaccinees^25^; we then explore a more general balance between history-based and polarized immunity: where the latter assumes a proportion of ‘fully’ immunized vaccinees at the start of the epidemic. We show that this heterogeneity can have a marked effect on the population impact of breakthrough infection; however, firm evidence of widespread polarized immunity against COVID-19 is lacking, so that the history-based assumption is probably reasonable initially.

We find that introducing heterogeneity in transmission into our basic model reduces the impact of the transmission interaction between susceptible and vaccinee subpopulations. This is mainly because population heterogeneity reduces average ℛ_0_; as illustrated by scaling ℛ_0_ to be constant at different levels of heterogeneity, which maintains the impact of vaccine breakthrough. For illustration, we focused on heterogeneities in mixing between vaccinated and susceptible populations; more complex patterns (eg adding high and low transmitters within these groups; results not shown) generate similar qualitative conclusions.

One arguably overoptimistic feature of our basic invasion model is that we assume a very highly vaccinated population at the start. In practice, there are likely to be significant pools of susceptibility to COVID-19 infection, for example associated with incidence of vaccine hesitancy or susceptibility in younger children. Even given standard spatial and social heterogeneities in mixing^30^, we cannot exclude significant mixing between susceptibles and vaccinees at some scales (for example parents and younger children). Estimating the impact of these heterogeneities on the broader epidemic is an urgent area for future work, as is the impact of waning and boosting of natural immunity.

One potential route to reduce vaccine breakthrough and its potential broader consequences is by administration of vaccine boosters^31^. However, this strategy only works in countries with access to ample vaccine supplies and the ability to administer multiple doses of complex vaccines over time. Without a major investment in equitable global mass vaccination, we cannot exclude the likelihood of significant future evolution of transmissive and immunologically elusive new variants^7^. Our simple invasion model illustrates a possible future for highly vaccinated countries in a world of vaccine nationalism: a series of new invading variants with ensuing epidemics and a scramble to revaccinate. The only way off this immunological rollercoaster is by a major and rapid investment in equitable global mass vaccination against SARS-CoV-2 and the development of second and third generation vaccines with enhanced transmission-blocking ability and increased duration of protection.

## Data Availability

This manuscript contains no data

## 1 Acknowledgments

This work was funded in part by the Natural Sciences and Engineering Research Council of Canada through a Postgraduate-Doctoral Scholarship (C.M.S.-R.); a Charlotte Elizabeth Procter Fellow-ship of Princeton University (C.M.S.-R.), the James S. McDonnell Foundation 21st Century Science Initiative Collaborative Award in Understanding Dynamic and Multi-scale Systems (S.A.L.); the C3.ai Digital Transformation Institute and Microsoft Corporation (S.A.L.); Gift from Google, LLC (S.A.L.); the National Science Foundation (CNS-2041952, CCF2142997, and CCF1917819) (S.A.L.); the US Centers for Disease Control and Prevention (B.T.G.); and the Flu Lab (B.T.G.).

