## Supplementary Information for "Vaccine breakthrough and the invasion dynamics of SARS-CoV-2 variants"

**4 Estimating $\epsilon$ from $VE$** **5**

**5 Supplementary figures** **5**

*Lewis-Sigler Institute for Integrative Genomics, Princeton University
†Department of Ecology and Evolutionary Biology, Princeton University
‡Department of Applied Mathematics and Theoretical Physics, University of Cambridge
§The Wellcome Trust, UK
¶Department of Bioengineering, McGill University
‖Princeton School of Public and International Affairs, Princeton University

### 1 Final size relation

#### 1.1 Basic model with a unique infection class

For Model (1), let $S_0 = S(0)$, $V_0 = V(0)$, $I_0 = I(0)$, $S_\infty = \lim_{t\to\infty} S(t)$, and $V_\infty = \lim_{t\to\infty} V(t)$. Using the matrix-theoretic approach [1], the final size relation is

$$-\ln\left(\frac{S_\infty}{S_0}\right) = \frac{\beta S_0}{\gamma}\left(1-\frac{S_\infty}{S_0}\right) + \frac{\beta V_0}{\gamma}\left(1-\left(\frac{S_\infty}{S_0}\right)^{\epsilon}\right) + \frac{\beta I_0}{\gamma}, \tag{S1a}$$

$$V_\infty = V_0\left(\frac{S_\infty}{S_0}\right)^{\epsilon}, \tag{S1b}$$

with the total fraction of infected individuals $\mathcal{I}_{\text{infected}} = S_0 - S_\infty + V_0 - V_\infty + I_0$.

#### 1.2 Model that distinguishes based on infection-type

Let $I_S^{(0)} = I_S(0)$ and $I_V^{(0)} = I_V(0)$. For Model (6), the final size relation becomes

$$-\ln\left(\frac{S_\infty}{S_0}\right) = \frac{\beta S_0}{\gamma}\left(1-\frac{S_\infty}{S_0}\right) + \frac{\beta V_0}{\gamma}\left(1-\left(\frac{S_\infty}{S_0}\right)^{\epsilon}\right) + \frac{\beta}{\gamma}({I_S}^{(0)} + {I_V}^{(0)}), \tag{S2a}$$

$$V_\infty = V_0\left(\frac{S_\infty}{S_0}\right)^{\epsilon}. \tag{S2b}$$

Using the above relations, Eq. (S2a) can be rewritten in terms of $\frac{V_\infty}{V_0}$:

$$-\frac{1}{\epsilon}\ln\left(\frac{V_\infty}{V_0}\right) = \frac{\beta S_0}{\gamma}\left(1-\left(\frac{V_\infty}{V_0}\right)^{\frac{1}{\epsilon}}\right) + \frac{\beta V_0}{\gamma}\left(1-\frac{V_\infty}{V_0}\right) + \frac{\beta}{\gamma}\left({I_S}^{(0)} + {I_V}^{(0)}\right), \tag{S3}$$

Since $\mathcal{R}_{0,I} = \frac{\beta}{\gamma}$, Eq. (S2a) becomes

$$-\ln\left(\frac{S_\infty}{S_0}\right) = \mathcal{R}_{0,I} S_0\left(1-\frac{S_\infty}{S_0}\right) + \mathcal{R}_{0,I} V_0\left(1-\left(\frac{S_\infty}{S_0}\right)^{\epsilon}\right) + \mathcal{R}_{0,I}({I_S}^{(0)} + {I_V}^{(0)}). \tag{S4}$$

Here and in all that follows, the fractions of individuals that are infected during the epidemic are given by

$$S_{\text{infected}} = S_0 - S_\infty + I_S^{(0)}, \tag{S5a}$$

$$V_{\text{infected}} = V_0 - V_\infty + I_V^{(0)}. \tag{S5b}$$

### 2 Immunological uncertainties

#### 2.1 Model with reduced transmission for breakthrough infections

##### 2.1.1 Formulation

If $I_V$ individuals individuals transmit at rate $\alpha\beta$, with $\alpha < 1$, then the model becomes

$$\frac{dS}{dt} = -\beta S(I_S + \alpha I_V), \tag{S6a}$$

$$\frac{dI_S}{dt} = \beta S(I_S + \alpha I_V) - \gamma I_S, \tag{S6b}$$

$$\frac{dV}{dt} = -\epsilon\beta V(I_S + \alpha I_V), \tag{S6c}$$

$$\frac{dI_V}{dt} = \epsilon\beta V(I_S + \alpha I_V) - \gamma I_V. \tag{S6d}$$

##### 2.1.2 Final size relation

For this model, the final size relation is then

$$-\ln\left(\frac{S_\infty}{S_0}\right) = \mathcal{R}_{0,I} S_0 \left(1 - \frac{S_\infty}{S_0}\right) + \mathcal{R}_{0,I}\alpha V_0 \left(1 - \left(\frac{S_\infty}{S_0}\right)^{\epsilon}\right) + \mathcal{R}_{0,I}({I_S}^{(0)} + \alpha {I_V}^{(0)}), \tag{S7}$$

with $V_\infty = V_0 \left(\frac{S_\infty}{S_0}\right)^{\epsilon}$.

#### 2.2 Mixed history-polarized immunity model

As described in the main text, we assume optimistically that a fraction $\epsilon_2$ of vaccinated individuals are fully immunized against infection. Thus, there are $V_0^* = (1 - \epsilon_2)V_0$ vaccinated individuals that are susceptible to breakthrough infection, with reduction in susceptibility $\epsilon_1$.

Thus, (S7) is now instead

$$-\ln\left(\frac{S_\infty}{S_0}\right) = \mathcal{R}_{0,I} S_0 \left(1 - \frac{S_\infty}{S_0}\right) + \mathcal{R}_{0,I}\alpha V_0^* \left(1 - \left(\frac{S_\infty}{S_0}\right)^{\epsilon_1}\right) + \mathcal{R}_{0,I}({I_S}^{(0)} + \alpha {I_V}^{(0)}), \tag{S8a}$$

$$V_\infty^* = V_0^* \left(\frac{S_\infty}{S_0}\right)^{\epsilon_1}. \tag{S8b}$$

and so $V_0^* - V_\infty^* + I_V^{(0)}$ vaccinated are infected during the epidemic, which corresponds to a fraction $\frac{V_0^* - V_\infty^* + I_V^{(0)}}{V_0 + I_V^{(0)}}$ of vaccinees.

### 3 Transmission heterogeneities

We next examine two situations of heterogeneity: one where the average transmission decreases because the across-group transmission decreases, and the other where this average transmission is kept constant.

#### 3.1 Decrease in average transmission

First, notice that by defining

$$G_1 = -\log S + \mathcal{R}_{0,I} S + \kappa \mathcal{R}_{0,I} V + \mathcal{R}_{0,I} I_S + \kappa \mathcal{R}_{0,I} I_V, \tag{S9a}$$

$$G_2 = -\log V + \epsilon\kappa \mathcal{R}_{0,I} S + \epsilon \mathcal{R}_{0,I} V + \epsilon \mathcal{R}_{0,I} \kappa I_S + \epsilon \mathcal{R}_{0,I} I_V, \tag{S9b}$$

then $G_1' = G_2' = 0$. Thus, in this case, the final size relations are

$$-\log \frac{S_\infty}{S_0} = \mathcal{R}_{0,I} S_0 \left(1 - \frac{S_\infty}{S_0}\right) + \kappa \mathcal{R}_{0,I} V_0 \left(1 - \frac{V_\infty}{V_0}\right) + \mathcal{R}_{0,I}(I_S^{(0)} + \kappa I_V^{(0)}) \tag{S10a}$$

$$-\log \frac{V_\infty}{V_0} = \epsilon\kappa \mathcal{R}_{0,I} S_0 \left(1 - \frac{S_\infty}{S_0}\right) + \epsilon \mathcal{R}_{0,I} V_0 \left(1 - \frac{V_\infty}{V_0}\right) + \epsilon \mathcal{R}_{0,I}(\kappa I_S^{(0)} + I_V^{(0)}) \tag{S10b}$$

#### 3.2 Constant average transmission

If, instead, transmission is normalized by $\frac{1+\kappa}{2}$ so as to keep the average transmission rate constant, then the Eqs (S10) become

$$-\log \frac{S_\infty}{S_0} = \frac{\mathcal{R}_{0,I}}{\frac{1+\kappa}{2}} S_0 \left(1 - \frac{S_\infty}{S_0}\right) + \frac{\kappa \mathcal{R}_{0,I}}{\frac{1+\kappa}{2}} V_0 \left(1 - \frac{V_\infty}{V_0}\right) + \frac{\mathcal{R}_{0,I}}{\frac{1+\kappa}{2}}(I_S^{(0)} + \kappa I_V^{(0)}) \tag{S11a}$$

$$-\log \frac{V_\infty}{V_0} = \frac{\epsilon\kappa \mathcal{R}_{0,I}}{\frac{1+\kappa}{2}} S_0 \left(1 - \frac{S_\infty}{S_0}\right) + \frac{\epsilon \mathcal{R}_{0,I}}{\frac{1+\kappa}{2}} V_0 \left(1 - \frac{V_\infty}{V_0}\right) + \frac{\epsilon \mathcal{R}_{0,I}}{\frac{1+\kappa}{2}}(\kappa I_S^{(0)} + I_V^{(0)}) \tag{S11b}$$

### 4 Estimating $\epsilon$ from $VE$

Using the basic model in the main text (Eqs. (2)), we can divide Eq. (2b) by Eq. (2a) to obtain

$$\frac{dV}{dS} = \epsilon V/S. \tag{S12}$$

Near the start of the epidemic ($t = 0$), when cases are rare, $S$ and $V$ are changing slowly, so that we can approximate the ratio $V/S$ by the constant $V(0)/S(0)$. Integrating $dV/dS$ between 0 and $t$ and re-arranging, we then obtain

$$\epsilon = \frac{(V(t) - V(0))/V(0)}{(S(t) - S(0))/S(0)}. \tag{S13}$$

Since $(V(0)-V(t))/V(0)$ and $(S(0)-S(t))/S(0)$ are the respective proportions of vaccinees and susceptibles infected, the right hand side of this equation is the basis of a standard vaccine risk ratio ($RR$) calculation [2]; $VE$ is then calculated as $VE = 1-RR = 1-\epsilon$. This illustrates the mapping of $1 - VE$ onto $\epsilon$ when cases are low and $V$ and $S$ are changing slowly.

To explore the more general case when $I$ is changing rapidly during the epidemic, we simulate the basic model for $\epsilon = 0.2$ (Figure S3). As predicted, $1 - VE$ approximates $\epsilon$ well when $I$ is small. However, during the peak of the epidemic, the figure shows that $1 - VE$ overestimates $\epsilon$; essentially, this is because $S$ is declining much more rapidly than $V$, shifting the ratio in equation (above). Thus, in general, $1 - VE$ gives if anything an upper bound for $\epsilon$. This is a simple case of the many subtleties in $VE$ calculations arising from epidemic dynamics [3].

### 5 Supplementary figures

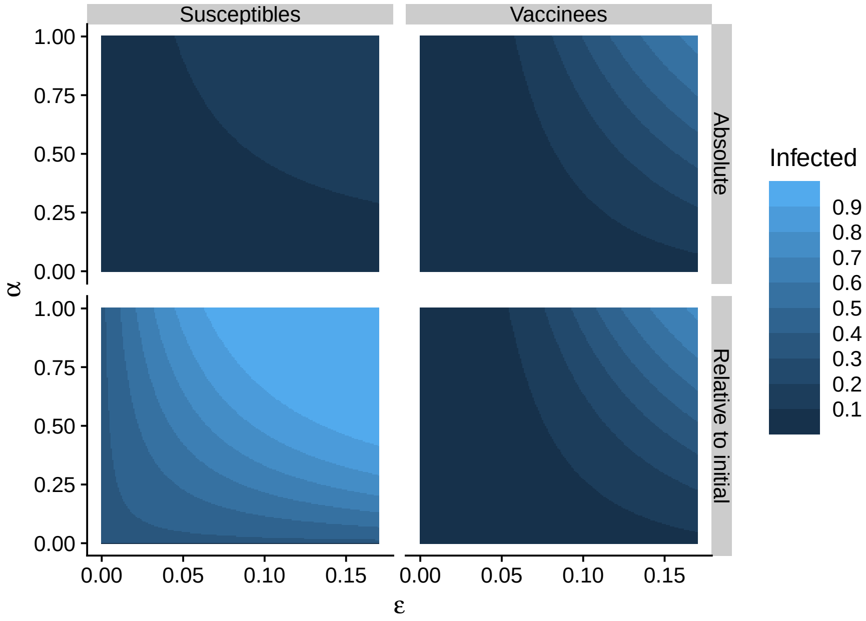

**Figure S1: Effect of $\alpha$ on model dynamics, with $\mathcal{R}_{0,I} = 10$. The sub-panels in this figure are as in Figure 2.**

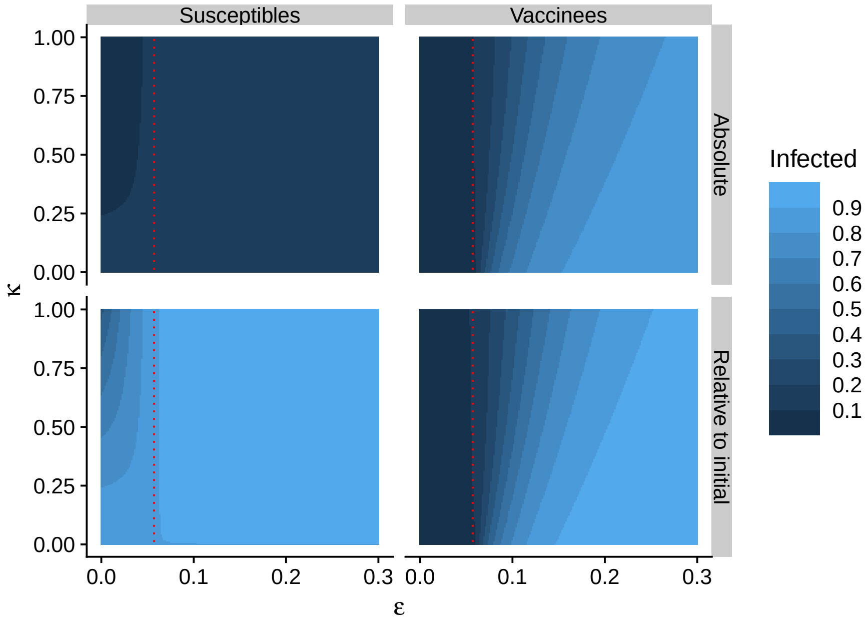

**Figure S2: Effect of normalized transmission on model dynamics, assuming $\kappa > 0$. This figure is as in Figure 4, but transmission is instead normalized by $\frac{1+\kappa}{2}$, so as to keep the average transmission constant.**

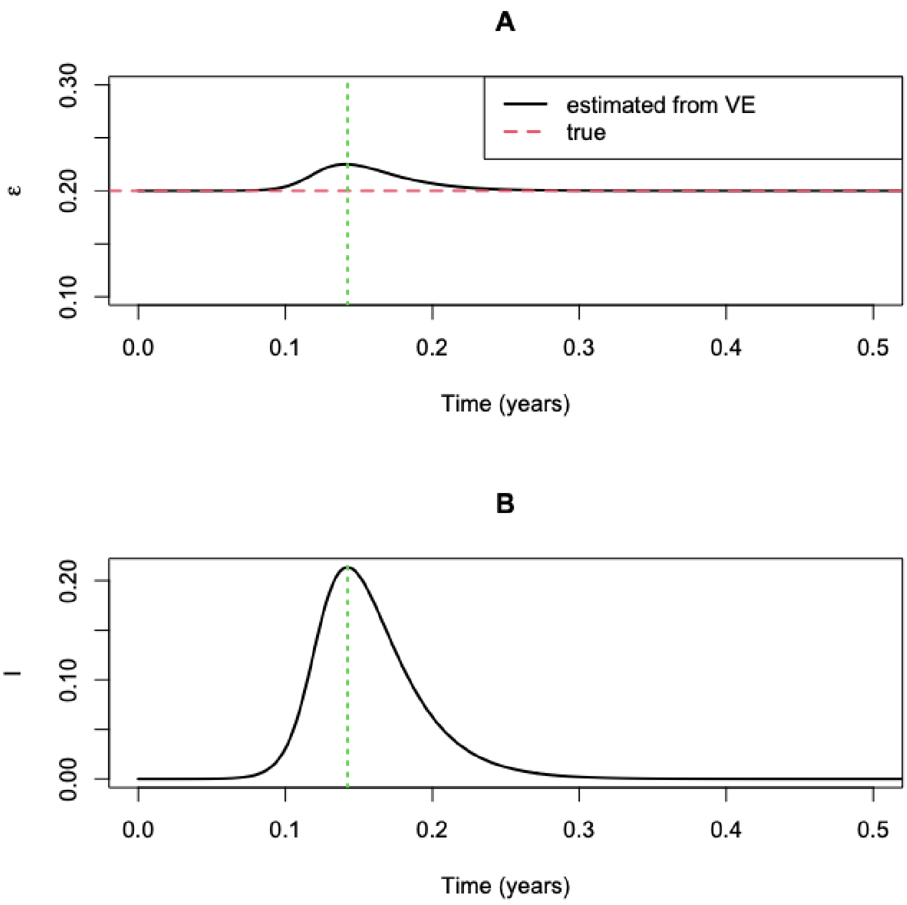

**Figure S3: Relationship between $\epsilon$ and $VE$ estimates. ($A$) Change in $\epsilon$ estimates from $VE$ over time during the epidemic. ($B$) Prevalence during the epidemic. In both panels, the green dashed line indicates the maximum prevalence (and corresponding estimate of $\epsilon$).**
